# High Resolution CHEST CT(HRCT) Evaluation in Patients Hospitalized with COVID-19 Infection

**DOI:** 10.1101/2020.05.26.20114082

**Authors:** Maulin Patel, Junad Chowdhury, Matthew Zheng, Osheen Abramian, Steven Verga, Huaqing Zhao, Nicole Patlakh, Nicholas Montecalvo, David Fleece, Gary Cohen, Maruti Kumaran, Chandra Dass, Gerard J. Criner, for the Temple University COVID-19 Research Group

## Abstract

**Introduction:** Currently the main diagnostic modality for COVID-19 (Coronavirus disease-2019) is reverse transcriptase polymerase chain reaction (RT-PCR) via nasopharyngeal swab which has high false negative rates. We evaluated the performance of high-resolution computed tomography (HRCT) imaging in the diagnosis of suspected COVID-19 infection compared to RT-PCR nasopharyngeal swab alone in patients hospitalized for suspected COVID-19 infection.

**Methods:** This was a retrospective analysis of 324 consecutive patients admitted to Temple University Hospital. All hospitalized patients who had RT-PCR testing and HRCT were included in the study. HRCTs were classified as Category 1, 2 or 3. Patients were then divided into four groups based on HRCT category and RT-PCR swab results for analysis.

**Results:** The average age of patients was 59.4 (±15.2) years and 123 (38.9%) were female. Predominant ethnicity was African American 148 (46.11%). 161 patients tested positive by RT-PCR, while 41 tested positive by HRCT. 167 (52.02%) had category 1 scan, 63 (19.63%) had category 2 scan and 91 (28.35%) had category 3 HRCT scans. There was substantial agreement between our radiologists for HRCT classification (κ = 0.64). Sensitivity and specificity of HRCT classification system was 77.6 and 73.7 respectively. Ferritin, LDH, AST and ALT were higher in Group 1 and D-dimers levels was higher in Group 3; differences however were not statistically significant.

**Conclusion:** Due to its high infectivity and asymptomatic transmission, until a highly sensitive and specific COVID-19 test is developed, HRCT should be incorporated into the assessment of patients who are hospitalized with suspected COVID-19.

**Key Points:** *Key Question:* Can High Resolution CT chest (HRCT) improve diagnostic accuracy of current Nasopharyngeal swab in suspected COVID-19 patients?

*Bottom Line:* In this retrospective analysis, our novel HRCT classification identified 20% of all COVID-19 patients who had negative nasopharyngeal reverse transcriptase polymerase chain reaction (RT-PCR) tests but had HRCT findings consistent with COVID-19 pneumonia. These patients were ruled out for other infections and laboratory markers were similar to other RT-PCR positive patients

*Why Read on:* Our new HRCT classification when combined with RT-PCR can improve diagnostic accuracy while promptly improving triaging in COVID-19 patients.

## Introduction

As of April 20th, 2020, Coronavirus disease 2019 (COVID-19) has affected 205 countries and territories with over 2.4 million confirmed cases.^1^ COVID-19 infection has created an influx of patients presenting for evaluation and treatment with acute and severe respiratory complaints- predominately dyspnea, fever, cough and hypoxemic respiratory failure. The severity and acuity of patient’s complaints coupled with the propensity for an abrupt decline in respiratory status necessitates an immediate diagnosis of COVID-19 infection and the ability to gauge the magnitude of pulmonary impairment. Unfortunately, conventional laboratory reverse transcriptase polymerase chain reaction (RT-PCR) testing of the nasopharynx/oropharynx has been fraught with delay, inefficiency, and suboptimal levels of sensitivity.^2,3^ Chest X-ray is limited in its ability to detect the presence and extent of lung involvement with COVID-19 infection.^4^ In contrast, high-resolution chest computed tomography (HRCT) has been reported to provide immediate results with a high sensitivity and specificity for detecting COVID-19 infection and the extent of lung involvement ^2,5,6^ However, the American College of Radiology has recommended against its routine use to detect COVID-19 infection^7^.

In preparation for the COVID-19 pandemic at our institution, we implemented a clinical care pathway whereby patients suspected of COVID-19 infection based on their risks and clinical presentation underwent RT-PCR based nasopharyngeal (NP) testing and HRCT imaging upon hospitalization. Herein, we report the clinical data and outcomes in patients who are COVID-19 NP swab positive or negative with their HRCT results in a large cohort of consecutively hospitalized patients suspected to have COVID-19 infection.

## Methods

### Temple University Hospital COVID-19 Care Pathway

Patients presenting with clinically suspicious COVID-19 infection were admitted to a dedicated COVID-19 hospital building specifically designed to avoid cross contamination with non-COVID-19 patients. Nasopharyngeal RT-PCR, HRCT, and relevant inflammatory markers were acquired prior to admission. Severity of disease was classified based on vital signs, oxygen requirements, and percentage of lung involvement on HRCT. Based on this classification, patients were screened for clinical trials targeting antiviral therapy and/or anti-inflammatory treatment.

### Patient population

The study consists of consecutive patients admitted to Temple University Hospital in Philadelphia, Pennsylvania, between March 10th, 2020, and April 6th, 2020. The study was derived from the IRB approved Temple University Registry for COVID19 (TUIRB Protocol Number: 26854). Subsequently, a separate IRB approval was granted for chart review of HRCT scans (TUIRB protocol number: 27051). A waiver of consent was granted due to minimal risk. All identifiable personal information was removed for privacy protection

During this period, we evaluated 324 consecutive patients with fevers or acute respiratory symptoms suggestive of COVID-19 infection that presented to the emergency room with severe symptoms that warranted hospitalization. Decisions to admit were primarily based on the severity of respiratory symptom burden and hypoxemia. Of these we included 321 patients for analysis who had both HRCT (1.25 mm slice thickness) and nasopharynx RT-PCR testing results during their hospitalization. We collected admission laboratory data which included Complete Blood Count with differential, Ferritin, Lactate Dehydrogenase (LDH), D-dimer, interleukin-6(IL-6) levels, aspartate aminotransferase (AST), and alanine aminotransferase (ALT) and C-Reactive Protein (CRP). Demographic parameters were compared across all groups included age, sex, comorbidities, body mass index, and smoking status (smoker, never smoker).

### Diagnosis of COVID-19

Nasopharyngeal swabs utilized for COVID-19 testing were processed by the Temple University Hospital microbiology laboratory with the Luminex Magpix (Vendor, Texas) NxTAG CoV for the detection and identification of SARS-CoV-2. The reagent used was supplied by cepheid (Vendor, California) on a Gene-expert platform. This test has been validated but under review by the Food and Drug Administration is pending. The turnaround time for this test is now approximately 6-8 hours but was several days in the early phases of the infection when our specimens were sent to city and state laboratories. For patients with multiple RT-PCR assays, COVID-19 was confirmed if a single PCR was positive.

### HRCT Imaging Performance

A dedicated CT scanner (Phillips Brilliant 256) was allocated for potential COVID-19 patients to reduce cross-contamination with uninfected patients. All images were obtained with patients in supine position, with slice thickness of 1.25 mm, kernel lung, pitch of 0.763, rotation time 0.33 seconds and radiation dose of 120 kilovoltage. If a patient had multiple CT scans during the admission, the scan performed closest to the timing of the RT-PCR was utilized for this analysis.

### HRCT Scoring of COVID-19 Pattern and Severity

Three attending radiologists (two thoracic radiologists and a third non-thoracic radiologist each with more than 20 years’ experience), blinded to RT-PCR results, interpreted the HRCT scans independently and classified them into one of the 3 categories: Category 1 – consistent with multifocal pneumonia; Category 2 – indeterminate for multifocal pneumonia; Category 3 – not consistent with multifocal pneumonia (figure 1). Category 1 scans demonstrated multiple lesions including ground glass opacities (GGOs), crazy-paving, patchy consolidations, nodules, nodules with halo, reverse halo/perilobular pattern irrespective of location and laterality. Category 3 patients had CT findings consistent with an alternate diagnosis (e.g. no active disease, congestive heart failure, lobar pneumonia, cavitary lesions, pleural effusion, and emphysema). Category 2 patients had indeterminate CT chest findings which did not fit criteria for either Category 1 or Category 3. The epidemiological history and clinical symptoms (fever, cough, dyspnea) were available to all three radiologists. Kappa Score was used to compare their interpretations for consistency and validation.

**Figure 1:**
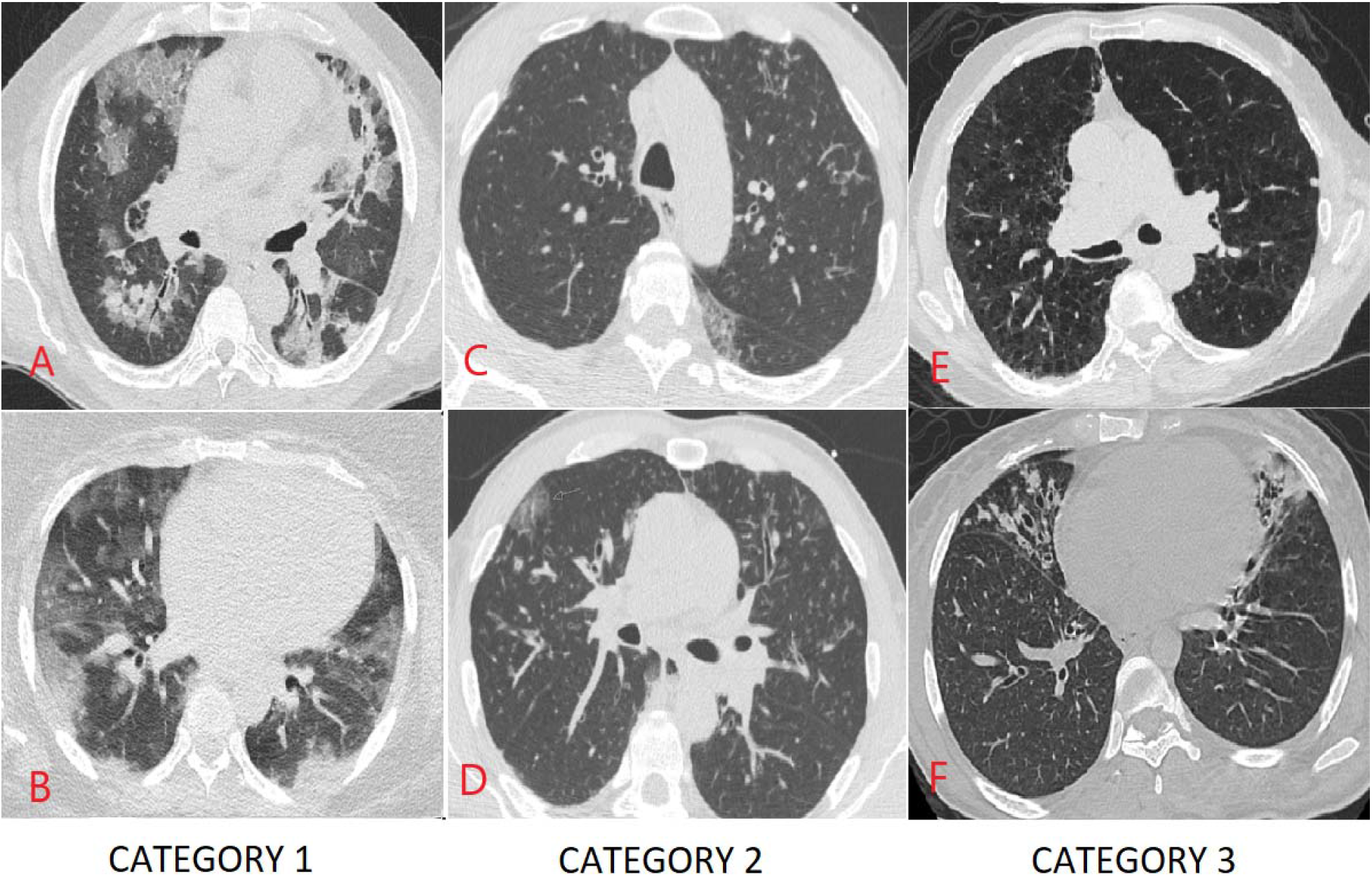
Examples of HRCT stratification system. Image A: Multifocal GGO, Crazy paving and lobar infiltrate. Image B: Multifocal GGO. Image C: Nodular Bronchiolitis. Image D: GGO with Bronchiectasis. Image E: Para-septal and centrilobular emphysema Image F: Bronchiectasis

### Treatments provided to patients

Patients with RT-PCR positive swabs were screened for eligibility for two randomized controlled trials involving sarilumab (Regeneron Pharmaceuticals; NCT04315298) and remdesivir (Gilead Sciences; NCT04292730 and NCT04292899). Those with significant disease who did not qualify for clinical trials were offered compassionate use with either anakinra or tocilizumab based on institutional care pathways. Other therapies included pulse-dose steroids (defined as a minimum daily 125 mg of methylprednisolone in any intermittent bolus frequency), intravenous immunoglobulin (IVIG), hydroxychloroquine (HCQ), and antibiotics. Therapies were offered based on clinical decompensation, worsening radiographic burden, and up trending inflammatory markers. These treatments were offered to all hospitalized patients with suspected COVID-19 infection based upon our institutional care plan.

### Statistical Methods

Continuous variables are presented as means (±SD), and categorical variables as numbers (%). Natural log transformation was employed to achieve normality for variables with skewed distribution. Continuous variables were compared with the use of the Analysis of Variance or two-sample t-test and categorical variables with the use of the Pearson chi-square test. The sensitivity, specificity, positive predictive value (PPV), negative predictive value (NPV), and accuracy of chest CT imaging was calculated using RT-PCR as reference. Receiver-operating-characteristic (ROC) curves were constructed to assess the sensitivity and specificity of chest CT imaging to compare their readings with PT-PCR results. All statistical tests were two-tailed, and P values of less than 0.05 were considered to indicate statistical significance. All statistical analyses were performed with the use of Stata 14.0 (StataCorp LP, College Station, TX).

For data analysis we divided patients into four groups:

Group 1: COVID (+) PCR and Category 1 CT scan

Group 2: COVID (+) PCR and Category 2 and 3 CT scan

Group 3: COVID (-) PCR and Category 1 CT scan

Group 4: COVID (-) PCR and Category 2 and 3 CT scan

Baseline demographics, clinical and laboratory parameters, as well as therapies offered were compared across four groups. We report the means and frequency of different variables across the four groups as well as sensitivity, specificity, negative predictive value, and positive predictive value of the new HRCT system. Laboratory markers were monitored for patients in Group 3 and compared to Group 1 and Group 3 independently.

## Results

### Patient population

321 patients were admitted to Temple University Hospital between March 10, 2020, and April 6, 2020 with suspected COVID-19 infection. The average age was 59.4 (±15.2) years and 123 (38.3%) were female. 148 (46.11%) were African American, 25(7.8%) were Caucasians, 64 (19.94%) were Hispanic, 29 (9.03%) were other races and 55 were unknown (17.1%). Major comorbidities included hypertension, diabetes, heart disease, lung disease and chronic kidney disease in decremental order. 26 of the 43 with Chronic kidney disease were also on hemodialysis. Patients in Group 4 were more likely to have underlying lung disease as a comorbid condition.

### Clinical/laboratory markers

Inflammatory markers including CRP, D-dimer, LDH, absolute lymphocyte count, AST and ALT are shown in Table 1. Inflammatory markers differed across the 4 groups. Patients in Group 1 and 3 had higher mean arterial pressures and temperatures. Ferritin, LDH, AST, and ALT were higher in Group 1 and D-dimers levels were higher in Group 3. IL-6 levels were highest in Group 2. Platelet counts were lower in Groups 1 and 2 while lymphocyte levels were highest in Group 4. Triglycerides were highest in Group 1. Overall, there was not a statistically significant difference in inflammatory marker levels between Groups 1 and 3 (Table 4).

**Table 1:** Demographics data including laboratory and clinical parameters across groups

|  | Group 1 | Group 2 | Group 3 | Group 4 | P-value |
| --- | --- | --- | --- | --- | --- |
| N | 125 (39.43%) | 36 (11.36%) | 41 (12.93%) | 115 (36.28%) |  |
| Age (yrs., Mean ± SD) | 57.5 (±14.8) | 61.6 (±17.2) | 57.3 (±12.5) | 61.4 (± 15.6) | 0.26 |
| Sex (F) n (%) | 46 (37.10 %) | 13 (36.11%) | 17 (41.45%) | 47 (40.87 %) | 0.89 |
| Race n (%) |  |  |  |  |  |
| African American | 59 (47.20%) | 20 (55.56%) | 18 (43.90%) | 51 (44.35%) | 0.260 |
| Caucasian | 6 (4.80%) | 2 (5.56%) | 6 (14.63%) | 11 (9.57%) |  |
| Hispanic | 25 (20%) | 6(16.67%) | 11(26.83%) | 21 (18.26%) |  |
| Other | 12 (9.60%) | 5 (13.89%) | 4 (9.76%) | 8 (6.96%) |  |
| BMI* kg/m2 (Mean ± SD) | 33.2 (±8.3) | 28.8 (±6.4) | 28.9 (±5.3) | 27.9 (±7.7) | 0.01 |
| Comorbidities n (%) |  |  |  |  |  |
| 1. Diabetes | 43 (44.79%) | 10 (33.3%) | 8 (28.6%) | 8 (40%) | 0.39 |
| 2. Hypertension | 64 (64.0%) | 19 (63.3%) | 20 (62.50%) | 15 (60%) | 0.98 |
| 3. Lung Dx | 28 (28.28 %) | 7 (23.3%) | 9 (28.1%) | 19 (76 %) | 0.001 |
| 4. Heart Dx | 24 (24.49%) | 9 (29.03%) | 7 (21.88%) | 9 (36%) | 0.61 |
| 5. Chronic kidney disease | 11 (10.48%) | 6 (18.18%) | 5 (12.82%) | 20 (21.98%) | 0.15 |
| Smoking^ n (%) |  |  |  |  |  |
| No | 58 (59.2%) | 12 (38.7%) | 15 (46.88%) | 9 (37.5%) |  |
| Yes | 40 (40.8%) | 19 (61.3%) | 17 (53.1%) | 15 (62.5%) | 0.09 |
| MAP* (Mean ± SD) | 95.5 (±16.0) | 87.1 (±17.5) | 94.9 (±18.5) | 90.3 (±20.2) | 0.01 |
| HR* (Mean ± SD) | 97.3 (±18.6) | 97.3 (±23.9) | 104.6(±20.1) | 95.4 (±20.0) | 0.083 |
| RR* (Mean ± SD) | 20.3 (±5.1) | 20.4(±5.8) | 20.8 (±4.8) | 20.3 (±4.6) | 0.35 |
| Temperature (Mean ± SD) | 99.7 (±1.6) | 99.2 (±2.8) | 99.4 (±2.2) | 98.7 (±1.6) | 0.001 |
| Pulse Oximetry (Mean ± SD) | 92.9 (±8.7) | 96.6 (±3.4) | 94 (±5.4) | 95.6 (±4.2) | 0.0001 |
| Laboratory Abnormalities (Mean ± SD) | | | | | |
| Ferritin (ng/ml) | 941 (±1462) | 555 (±602) | 392 (±385) | 396 (±1056) | 0.001 |
| CRP* (mg/dl) | 8.3 (±6.5) | 5.02 (±4.8) | 10.3 (±8.6) | 6.7 (±8.8) | 0.001 |
| LDH* (U/L) | 371.9(±271) | 253.45(±100) | 312.4 (±151) | 256.9 (±130) | 0.001 |
| D-Dimer (ng/ml) | 3150.3 (±10983) | 2722.3(±4213.6) | 4833(±14282) | 2972.4(±5771.5) | 0.184 |
| Fibrinogen (mg/dl) | 499.4 (±181.7) | 345.5 (±122) | 574.2(±207.3) | 399.4(±220.6) | 0.001 |
| Lym Count* | 1.1 (±0.53) | 1.2 (±0.68) | 1.1 (±0.74) | 1.6 (±1.1) | 0.001 |
| (K/mm<sup>3</sup>) |  |  |  |  |  |
| IL-6* (pg/ml) | 87.7 (±286.4) | 180.1(±265.9) | 86.6 (±111.6) | 31.5 (±47.5) | 0.001 |
| AST* (U/L) | 52.5 (±69.1) | 50.5 (±50.9) | 49.7 (±82.6) | 36.4 (±52.1) | 0.001 |
| ALT* (U/L) | 45.04 (±40.8) | 42.7 (±39.9) | 35.8 (±26.6) | 36.4 (±48.07) | 0.001 |
| Platelets (k/mm<sup>3</sup>) | 207.03 (±96.5) | 191.7 (±64.9) | 237.8 (±101.5) | 259.4 (±89.1) | 0.04 |
| Triglycerides | 249.42(±358.62) | 115 (±47.96) | 114.2(±61.5) | 104(±56.9) | 0.001 |

\*BMI - body mass index, MAP - Mean arterial pressure, HR – heart rate, RR – respiratory rate,
CRP =C-reactive protein, LDH - Lactate dehydrogenase, IL-6 - interleukin 6, AST -Aspartate
Aminotransferase, ALT - Alanine Aminotransferase, Lymph count - absolute lymphocyte count.
^smoking - current vs former/nonsmokers

### Level of agreement with Radiology interpretation

The Kappa statistic of 0.64 indicates substantial agreement between the three radiologists for the HRCT COVID-19 classification system. The consistency was highest for Category 1 (**K** = 0.79) and Category 3 (**K** = 0.69) HRCT scans, while Category 2 had the least agreement (**K** = 0.28). Figure 1 shows receiver-operating-characteristic (ROC) curves among three radiologists to assess the sensitivity and specificity of HRCT readings with PT-PCR results. The diagnostic accuracy of PCR + was substantial and consistent among our three radiologists (ROC area ranging from 0.7552 to 0.7691, figure 2).

**Figure 2:**
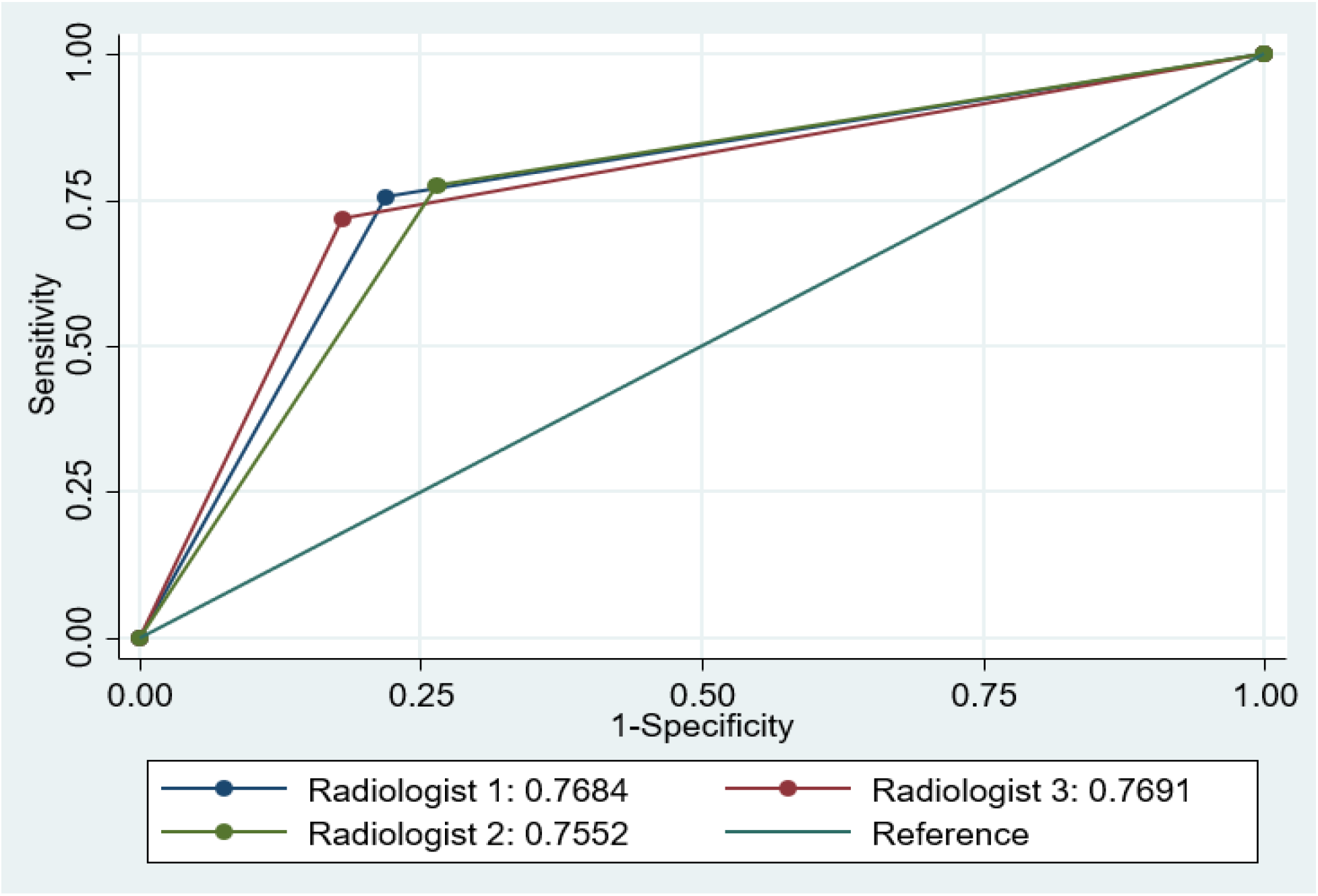
Receiver-operating-characteristic (ROC) curves among three radiologists

### Breakdown of patients into different categories based on imaging

A total of 161 patients tested positive by RT-PCR, while 156 were negative. 167 patients (52.02%) had Category 1 HRCT scans, 63 (19.63 %) had Category 2 scans and 91 (28.3%) had Category 3 scans (Table 2). Demographics and distribution of patients stratified between Group 1 to 4 is demonstrated in Table 1. Sensitivity and specificity of HRCT was 77.6% (95% CI: 70.4 - 83.8) and 73.7 (95% CI: 66.1 – 80.4) respectively. The positive predictive value and negative predictive values of HRCT for COVID-19 were calculated at 75.3% (95% CI: 68% - 81.7%) and 76.2% (95% CI: 68.6% - 82.7%) respectively. Overall, 41 patients were diagnosed via HRCT and 168 via RT-PCR swabs. Thus, 20% of total COVID cases that were declared negative by RT-PCR swab had characteristic findings of COVID-19 infection by HRCT.

**Table 2:** Frequency of each CT category with positive/negative PCR results

|  |  | COVID-PCR Result |  |
| --- | --- | --- | --- |
|  | Total | Positive | Negative |
| CT Category 1 | 167 (52.02%) | 125 (38.9 %) | 41 (12.8%) |
| CT Category 2 | 63 (19.63%) | 19 (5.9 %) | 42 (13.1%) |
| CT Category 3 | 91 (28.35%) | 17 (5.2%) | 73 (22.74 %) |

**Table 3:** Treatment distribution across groups

| Drug Therapy | Group 1 (n = 125) | Group 2 (n =36) | Group 3 (n = 41) | Group 4 (n=115) |
| --- | --- | --- | --- | --- |
| Randomized to Remdesivir trial | 23 (18.4%) | 2 (5.6%) | 0 | 0 |
| Randomized to Sarilumab trial | 37 (29.6%) | 2 (5.6 %) | 0 | 0 |
| Anakinra | 3 (2.4%) | 0 | 1 (2.4%) | 0 |
| Tocilizumab | 2 (1.6%) | 0 | 1 (2.4%) | 0 |
| IVIg | 12 (9.6%) | 0 | 5 (12.2%) | 0 |
| Pulse Steroids | 31 (24.8%) | 1 (2.7%) | 7 (17.07%) | 0 |
| Hydroxychloroquine | 11 (8.8%) | 1 (2.7%) | 5 (12.1%) | 0 |
| Azithromycin | 66 (52.8%) | 14 (38.9%) | 20 (48.8%) | 26 (22.6%) |
| Other Antibiotics | 63 (50.4%) | 16 (44.4%) | 22 (53.66%) | 36 (31.3%) |

### Breakdown of patients into different groups based on COVID-19 diagnosis by nasopharyngeal swab and imaging results

38.9 % (n=125) patients were classified as Group 1 (COVID (+) PCR and Category 1 CT scan); 11.2% (n=36) as Group 2 (COVID (+) PCR and Category 2 and 3 CT scan), 12.8% (n=41) as Group 3 (COVID (-) PCR and Category 1 CT scan) and 34.8 % (n=115) as Group 4 (COVID (-) PCR and Category 2 and 3 CT scan). In group 3, 38 (92.7 %) patients received respiratory viral panels (RVP) which were all negative; 21(51.2%) had sputum cultures, of which only 1 was positive for staph aureus. Repeat RT-PCR NP swab was performed in 28(68.3%) patients. which were all negative. When comparing Group 1 to Group 3, there were no statistically significant differences. Group 1 did have higher Ferritin and triglyceride levels, whereas Group 3 had higher CRP and D-dimer levels (Table 4). When comparing Groups 3 and 4, CRP, D-dimer, IL-6 was significantly higher, while lymphocyte count was significantly lower in Group 3.

**Table 4:** Two sample t-test of laboratory markers and treatments amongst RT-PCR positive patients (Group 1) and HRCT positive but PCR negative patients (Group 3).

|  | Group 1 (n =125) | Group 3 (n = 41) | p-value |
| --- | --- | --- | --- |
| Laboratory markers |  |  |  |
| Ferritin (ng/ml) | 941 (±1462) | 392 (±385) | 0.06 |
| CRP* (mg/dl) | 8.3 (±6.5) | 10.3 (±8.6) | 0.93 |
| LDH (U/L) | 371.9(±271) | 312.4 (±151) | 0.83 |
| D-Dimer (ng/ml) | 3150.3 (±10983) | 4833 (±14282) | 0.9 |
| Fibrinogen (mg/dl) | 499.4 (±181.7) | 574.2(±207.3) | 0.41 |
| Lymphocyte Count<br>(K/mm<sup>3</sup>) | 1.1 (±0.53) | 1.1 (±0.74) | 0.9 |
| IL- 6 (pg/ml) | 87.7 (±286.4) | 86.6 (±111.6) | 0.9 |
| AST (U/L) | 52.5 (±69.1) | 49.7 (±82.6) | 0.9 |
| ALT (U/L) | 45.04 (±40.8) | 35.8 (±26.6) | 0.9 |
| Platelets (k/mm3) | 207.03 (±96.5) | 237.8 (±101.5) | 0.39 |
| Triglycerides | 249.42(±358.62) | 114.2(±61.5) | 0.09 |
| Treatments |  |  |  |
| Remdesivir | 23 (18.4%) | 0 | NA* |
| Sarilumab | 37 (29.6%) | 0 | NA |
| Anakinra | 3 (2.4%) | 1 (2.4%) | 0.81 |
| Tocilizumab | 2 (1.6%) | 1 (2.4%) | 0.57 |
| IVIG | 12 (9.6%) | 5 (12.2%) | 0.33 |
| Pulse Steroids | 31 (24.8%) | 7 (17.07%) | 0.69 |
| Hydroxychloroquine | 11 (8.8%) | 5 (12.1%) | NA |
| Azithromycin | 66 (52.8%) | 20 (48.8%) | 0.89 |
| Other Antibiotics | 63 (50.4%) | 22 (53.66%) | 0.60 |

\*CRP - C-reactive protein, LDH - Lactate dehydrogenase, IL-6 - Interleukin - 6, AST - Aspartate Aminotransferase, ALT - Alanine Aminotransferase. NA - not applicable

**Table 5:** Two sample t-test of laboratory markers and treatment amongst RT-PCR neg patients, HRCT positive (Group 3) and HRCT/ PCR negative patients (Group 4).

|  | Group 3 (n = 41) | Group 4 (n=115) | p-value |
| --- | --- | --- | --- |

| Laboratory markers |  |  |  |
| --- | --- | --- | --- |
| Ferritin (ng/ml) | 392 (±385) | 396 (±1056) | 0.9 |
| CRP* (mg/dl) | 10.8 (±8.9) | 6.7 (±8.8) | 0.1 |
| LDH (U/L) | 312.4 (±151) | 256.9 (±130) | 0.9 |
| D-Dimer (ng/ml) | 6915.4 (±18822) | 2972.4(±5771.5) | 0.41 |
| Fibrinogen (mg/dl) | 574.2(±207.3) | 399.4(±220.6) | 0.9 |
| Lymphocyte Count<br>(K/mm<sup>3</sup>) | 1.1 (±0.74) | 1.6 (±1.1) | 0.04 |
| IL- 6 (pg/ml) | 86.6 (±111.6) | 31.5 (±47.5) | 0.001 |
| AST (U/L) | 49.7 (±82.6) | 36.4 (±52.1) | 0.9 |
| ALT (U/L) | 35.8 (±26.6) | 36.4 (±48.07) | 0.9 |
| Platelets (k/mm<sup>3</sup>) | 237.8 (±101.5) | 259.4 (±89.1) | 0.9 |
| Triglycerides | 114.2(±61.5) | 104(±56.9) | 0.9 |
| Treatments |  |  |  |
| Remdesivir | 0 | 0 | NA |
| Sarilumab | 0 | 0 | NA |
| Anakinra | 1 (2.4%) | 0 | 0.09 |
| Tocilizumab | 1 (2.4%) | 0 | 0.09 |
| IVIG | 5 (12.2%) | 0 | 0.0001 |
| Pulse Steroids | 7 (17.07%) | 2 (1.74%) | 0.0001 |
| Hydroxychloroquine | 5 (12.1%) | 0 | NA |
| Azithromycin | 20 (48.8%) | 26 (23%) | 0.002 |
| Other Antibiotics | 22 (53.66%) | 36 (31.3%) | 0.010 |

### Treatments/LOS by category

There were no statistically significant differences amongst Groups 1 and 3 with respect to treatments (Table 1). At the time of this analysis, 57.9% of Group 1, 61.1% of Group 2, 65% of Group 3 and 6 3% of Group 4 patients were discharged from the hospital. Mean length of stay for RT-PCR positive group (Groups 1 and 2) was 5.9(±4.3) days and 4.44(±2.97) days for Group 3 (p = 0.11).

## Discussion

Our data shows that 20% of patients admitted to the hospital with suspected COVID-19 have negative nasopharyngeal RT-PCR tests but have findings on HRCT consistent with COVID-19 infection. Furthermore, these patients demonstrated clinical signs and laboratory markers that were congruent with systemic inflammation. Nonsignificant higher levels of inflammatory markers in Group 1 is potentially related to factors contributing to severity of illness and the progression of hyperinflammatory syndromes characterized by cytokine release storms and secondary hemophagocytic lymphohistiocytosis^8^. In lieu of more sensitive COVID-19 testing, we conclude that HRCT should be utilized for the assessment and management of patients with suspected COVID-19 infection. HRCT, when considered in conjunction with inflammatory markers, can provide immediate aid to triage and initiate treatment planning.

Currently, nasopharyngeal (NP) and oropharyngeal (OP) swabs evaluated via RT-PCR are the tests of choice for diagnosing COVID-19. Literature suggests the sensitivity of the NP and OP RT-PCR is as low as 60%.^2^ 68.2% patients in Group 3 did have repeat RT-PCR swabs, however the turnaround time was usually 24-48 hours and all of them were negative. SARS-CoV-2 replication in the upper respiratory tract peaks in the first week post infection. After the first week, the positive predictive value of NP RT-PCR is worse when patients present with predominantly lower respiratory symptoms.^9,10^ Throat swab samples may increase the sensitivity at this stage of infection, but acquiring lower respiratory samples through tracheal aspirates or bronchoalveolar lavage poses a high risk of aerosolization and transmission to healthcare workers.^3,11^ False negative testing also risks inadvertent contamination of hospital wards sequestered for non-COVID-19 patients. Moreover, the current methods of diagnosis with nuclear extraction techniques take hours to days and further compounds the delay of diagnosis, triage, and implementation of appropriate therapy. Repeat testing to confirm positive or negative status further hinders timely clinical decision making. The prevalence of other viruses that might change treatment plans (i.e., influenza) was extremely low in our geographic region during this period. In addition, 92% of our group 3 patients had negative respiratory viral panel. Besides, the prevalence of severe respiratory failure with other viruses such as rhinovirus and other coronaviruses is temporally extremely low. When able to produce sputum, bacterial pneumonia was routinely screened with sputum cultures (51.2% of group 3) in our cohort; and in certain clinical scenarios the value of HRCT was augmented when dense lobar consolidations suggested bacterial pneumonia with prompt initiation of antibacterial treatment.

Our imaging stratification system shows that these deficiencies in laboratory testing can be mitigated somewhat by performing HRCT on admission. While RT-PCR positive patients in Groups 1 and 2 were definitively managed as COVID-19, Group 3 patients were also treated as active COVID-19 infection if clinical and laboratory presentation was consistent in the absence of strong consideration of an alternative diagnosis. Our HRCT stratification scheme had ~75% sensitivity and specificity in correlating with PCR findings. Despite a lower sensitivity, we report a higher specificity than that reported in literature.^2,11,12^ The slightly lower sensitivity may be explained by the radiologists’ assignment of uncommon but non-pathognomonic COVID-19 related HRCT findings that are more traditionally consistent with alternate diagnoses to categories 2 and 3. This system highlights the role of HRCTs in reducing the 30-40% false negative rate of RT-PCR testing. HRCT can thus help mitigate transmission in health care workers by identifying swab negative patients, thus reducing cost and burden on health care systems.

HRCT has been successfully used in China as a diagnostic modality when widespread RT-PCR was not available. In China, like our findings, 22% of the suspected initial COVID-19 cases were treated as presumptive COVID-19 without RT-PCR testing based on radiographic patterns, inflammatory markers, and clinical symptoms which is similar to our HRCT stratification system.^13^ About 80% of early HRCT findings range from isolated focal GGOs to multifocal GGO with a predominantly peripheral and peribronchovascular distribution. These opacities progress to consolidations or crazing paving (GGO with interlobular and intralobular septal thickening) in mid to late disease.^14-16^ Rarely, dense consolidation and reverse halo sign suggestive of organizing pneumonia has also been reported.^9,16,17^ Hence, HRCT as a diagnostic tool not only gives rapid and valuable information in suspected COVID-19 patients for appropriate triaging isolation, and treatment procedures, but also provides information on the extent of lung involvement thereby gauging risk of rapid respiratory decline. We do understand that performing HRCT for every suspect upper respiratory tract infection, flu-like illness, or pneumonia is not cost-effective nor sustainable in the long-term. This protocol is designed specifically for this time period, where reliable laboratory testing for COVID-19 leaves much to be desired. When very high sensitivity laboratory testing, such as rapid antibody testing, is developed and widely utilized, then HRCT will only be needed in select clinical scenarios.

Our study has several limitations. The most important is that we provide no measure of viral loads or antibody test results to confirm the absolute presence or absence of COVID-19 infection due to lack of availability. Not all clinical and laboratory data was available for all patients. Additionally, due to limited use of bronchoscopy out of aerosolization and contamination concerns, confirmatory bronchoalveolar lavage could not be performed to definitively stratify Group 3 patients as true false negatives. Interpretation of our findings might be limited by the sample size and should be validated in larger studies.

In conclusion, the findings outlined in this study suggests that a false negative RT-PCR population can be isolated for clinical re-evaluation if concomitant HRCT findings are consistent with a Category 1 stratification. We believe that combining RT-PCR with HRCT evaluation can increase the sensitivity and specificity of diagnostic testing to greater than 90%. To our knowledge, this is the first study that evaluates the utility of an HRCT based stratification system for the diagnosis of COVID-19. It is our contention that utilization of HRCT in the evaluation of a suspected COVID-19 infection will curb the false negativity of sole RT-PCR testing and help influence triage and clinical management decisions.

## Data Availability

All deidentified data will be available upon request to the correnponding author

## Acknowledgements/Author Contributions

Maulin Patel will be the corresponding author and gurantor for the manuscript. Maulin Patel, Gerard J Criner formulated the overall study design. Gerard J Criner, Gary Cohen, Maruti Kumaran and Chandra Dass designed the new HRCT system. Huaqing Zhao, Nicole Patlakh, David Fleece, Nicholas Montecalvo, Chandra Dass, Maruti Kumaran, Gary Cohen, Maulin Patel assisted in data collection, consolidation and analysis. Maulin Patel, Junad Chowdhury, Matthew Zheng, Osheen Abramian, Steven Verga drafted the manuscript. Gerard J Criner revised and reviewed the Manuscript

